# COVID-19 Vaccination Prioritization Based on Cardiovascular Risk Factors and Number-Needed-to-Vaccinate to Prevent Death

**DOI:** 10.1101/2021.03.24.21254227

**Authors:** Darryl P. Leong, Amitava Banerjee, Salim Yusuf

**Author notes:** **Corresponding author:** Darryl Leong. **Authors’ declaration:** The authors have no competing interests to declare.

## Abstract

The supply limitations of COVID-19 vaccines have led to the need to prioritize vaccine distribution. Obesity, diabetes and hypertension have been associated with an increased risk of severe COVID-19 infection. Approximately half as many individuals with a cardiovascular risk factor need to be vaccinated against COVID-19 to prevent related death as compared with individuals without a risk factor. Our analysis suggests that prioritizing adults with these cardiovascular risk factors for vaccination is likely to be an efficient way to reduce population COVID-19 mortality.

## INTRODUCTION

Vaccination is highly efficacious at preventing symptomatic COVID-19 infection(1-3). There has consequently been unprecedented global demand for effective COVID-19 vaccines, which has exceeded supply. Policymakers have implemented guidance for prioritising vaccine distribution in their countries and jurisdictions. These policies are designed to vaccinate those at highest risk of contracting COVID-19, being hospitalized or dying from the condition. Current policy prioritizes the elderly, especially those institutionalized, with evolving guidelines for other groups. The US Centers for Disease Control and Public Health England suggest that individuals with certain underlying health conditions, which are thought to be associated with increased COVID-19 morbidity and mortality, should be prioritized for vaccination. As of February 23, 2021, Public Health England’s guidance for vaccination is that adults ≥65 years be prioritized highest; followed by adults <65 years with diabetes, body-mass index ≥40kg/m^2^, or other chronic disease; with healthy adults <65 years prioritized lowest. However, the public health benefits of such guidance have not been demonstrated and in the absence of data on the population distribution of these morbidities, the value of the strategies proposed cannot be quantified and are speculative.

Younger age and male sex are associated with an increased risk of acquiring COVID-19, while in those who develop infection, older age and male sex are associated with a higher risk of death. Cardiovascular risk factors are recognized risk factors for acquiring COVID-19 (obesity and diabetes)(4) and have also been associated with a higher case-fatality rate among those developing COVID-19 (obesity, diabetes and hypertension)(5, 6). Therefore, adults in the general population with cardiovascular risk factors are likely to be at higher risk of acquiring COVID-19 and also having a higher mortality should they get infected. Consequently, the absolute reduction in COVID-19 risk from vaccinating these individuals might be expected to be higher than non-obese individuals without diabetes or hypertension. The objective of this analysis was to estimate the number of middle-aged and older adults (age 40-80 years) needed-to-vaccinate to prevent a COVID-19 death in populations with different clinical characteristics.

## DATA SOURCES

The numbers of men and women stratified by age in Canada in 2020 were obtained from estimates from the United Nations Department of Economic and Social Affairs(7). The Canadian incidence rates of COVID-19 for the week of 31 January to 6 February, 2021, were obtained from the Government of Canada’s website on COVID-19 epidemiological and economic research data(8). We estimated the age-stratified and overall prevalence rates of obesity, diabetes and hypertension using data from the Prospective Urban Rural Epidemiology (PURE) study – a large, prospective cohort study including 10,462 adults from British Columbia, Ontario and Quebec. Obesity was defined as a body-mass index ≥30kg/m^2^. Diabetes included self-reported diabetes, use of blood glucose-lowering medications or a fasting blood glucose level ≥7mmol/L. Hypertension included self-reported hypertension, use of a blood pressure-lowering medication or blood pressure ≥140/90mmHg(9). We observed a relative risk for COVID-19 infection among obese individuals (as compared with those with a body-mass index 20 to <30kg/m^2^) of 1.53; a relative risk for COVID-19 infection among adults with diabetes of 1.50; and a relative risk for COVID-19 infection among adults with hypertension of 1.17 based on our analysis of 10,090 individuals from the PURE data (unpublished). We used Public Health Ontario’s estimates of the COVID-19 case-fatality ratio, which included data up to May, 2020(10). The effects of obesity on COVID-19 case-fatality rates were estimated using data from a systematic review in which BMI ≥30kg/m^2^ was associated with an odds ratio for death of 1.67 (95% CI 1.43-1.96)(11). Based on another systematic review of the effects of co-morbidities on COVID-19 outcomes, we assumed that those with diabetes and COVID-19 had a relative risk of death of 1.94 and that those with hypertension and COVID-19 had a relative risk of death of 2.10 as compared with infected individuals without diabetes or hypertension respectively(5). We estimated the protective effect of vaccination by pooling (using random effects models) the estimates from three randomised trials evaluating the Pfizer, Moderna and Oxford-Astra Zeneca COVID-19 vaccines respectively(1-3).

We estimated the number-needed-to-vaccinate in each stratum of the Canadian Population as 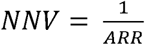 where the absolute risk reduction,

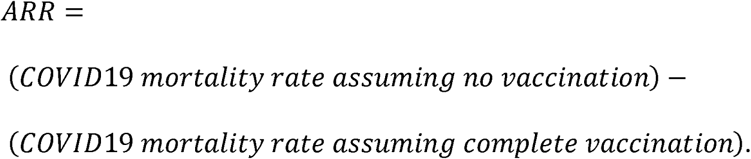

## FINDINGS

In Canada, in 2020, there were 2,414,000 men aged 40-50 years; 2,597,000 men aged 50-60 years; 2,322,000 men aged 60-70 years; and 1,435,000 men aged 70-80 years. The respective numbers of women in these age categories were 2,433,000; 2,585,000; 2,390,000; and 1,583,000. In Canada, the incidence rate of COVID-19 per 100,000 people from 31 January to 6 February, 2021, was 85.2 among men aged 40-50 years; 72.7 among men aged 50-60 years; 53.1 among men aged 60-70 years; and 40.3 among men aged 70-80 years. Respective rates among women were 84.5; 71.3; 44.4; and 39.2. In the PURE study, rates of obesity, diabetes and hypertension among Canadian participants (above 35 years of age) were respectively 26%, 9% and 38%. Rates of these risk factors stratified by age and sex are presented in the Table. The pooled effect of COVID-19 vaccination on the risk of acquiring COVID-19 (Figure 1) was a relative risk (95% CI) of 0.10 (0.03-0.34). Among those who are infected with COVID-19, the case-fatality ratio (i.e. the proportion of identified cases that succumb to the infection adjusted for censoring bias) according to Public Health Ontario data is 0.67% for those aged 40-50 years; 2.03% for those aged 50-60 years; 6.52% for those aged 60-70 years; and 20.89% for those aged 70-80 years. Using these data, we estimated the numbers of individuals 1) in each age, sex and body-mass index stratum; 2) among those with and without diabetes; and 3) among those with and without hypertension who would develop and die from COVID-19 in the absence of vaccination versus the numbers expected to develop and die from COVID-19 following vaccination. Based on these data, the estimated numbers-needed-to vaccinate to prevent one COVID-19 death in Canadian adults overall is 33,595 and in men aged 70-80 years is 8722 and in women aged 70-80 years is 9060. The estimated numbers-needed-to vaccinate to prevent one COVID-19 death in different subgroups of the adult population are presented in the Figure 2.

**Table.**
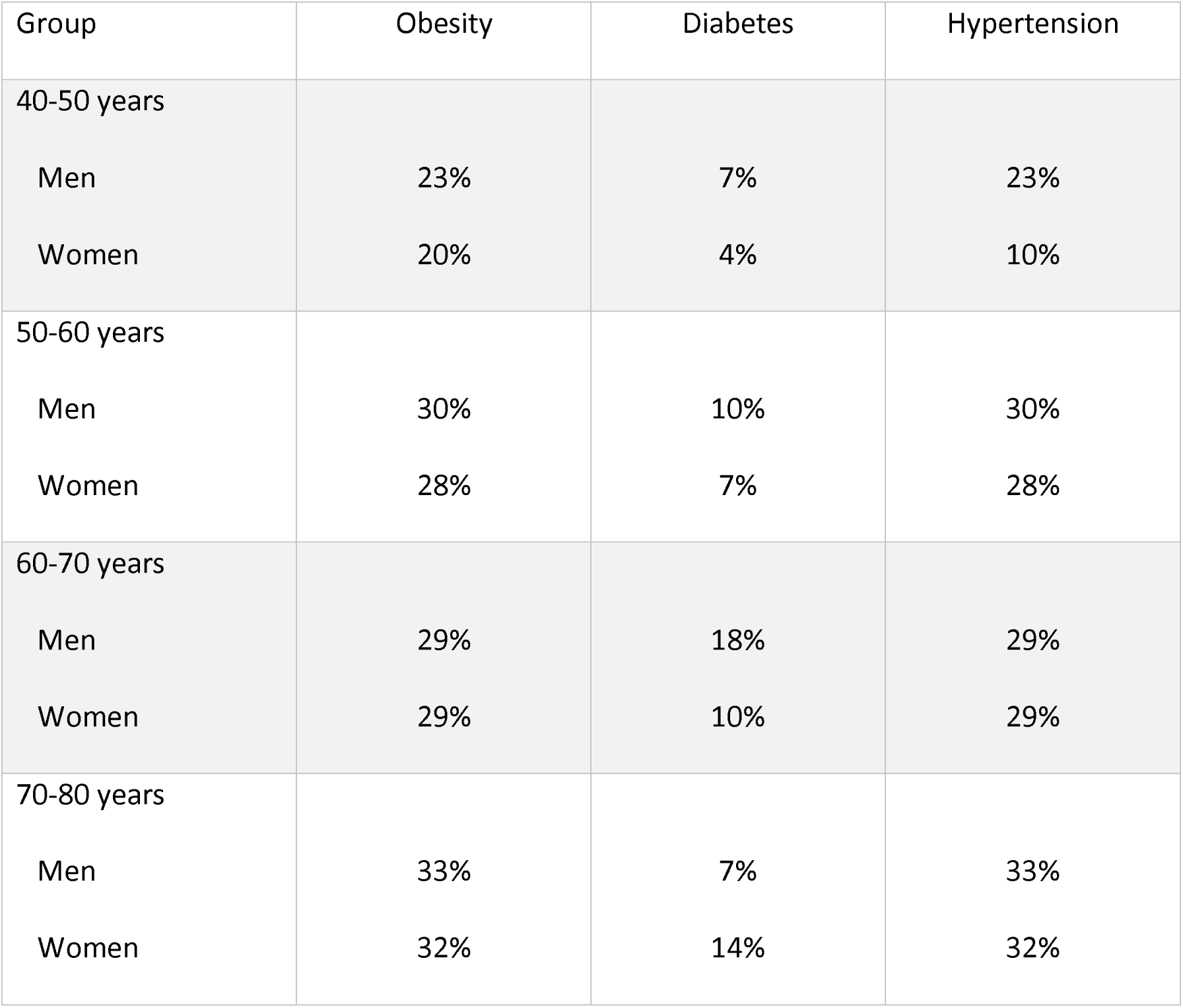
Prevalence rates of obesity, diabetes and hypertension among 11,000 Canadian participants aged 35 to 70 years in the Prospective Urban Rural Epidemiology (PURE) study.

| Group | Obesity | Diabetes | Hypertension |
| --- | --- | --- | --- |
| 40-50 years |  |  |  |
| Men | 23% | 7% | 23% |
| Women | 20% | 4% | 10% |
| 50-60 years |  |  |  |
| Men | 30% | 10% | 30% |
| Women | 28% | 7% | 28% |
| 60-70 years |  |  |  |
| Men | 29% | 18% | 29% |
| Women | 29% | 10% | 29% |
| 70-80 years |  |  |  |
| Men | 33% | 7% | 33% |
| Women | 32% | 14% | 32% |

**Figure 1.**
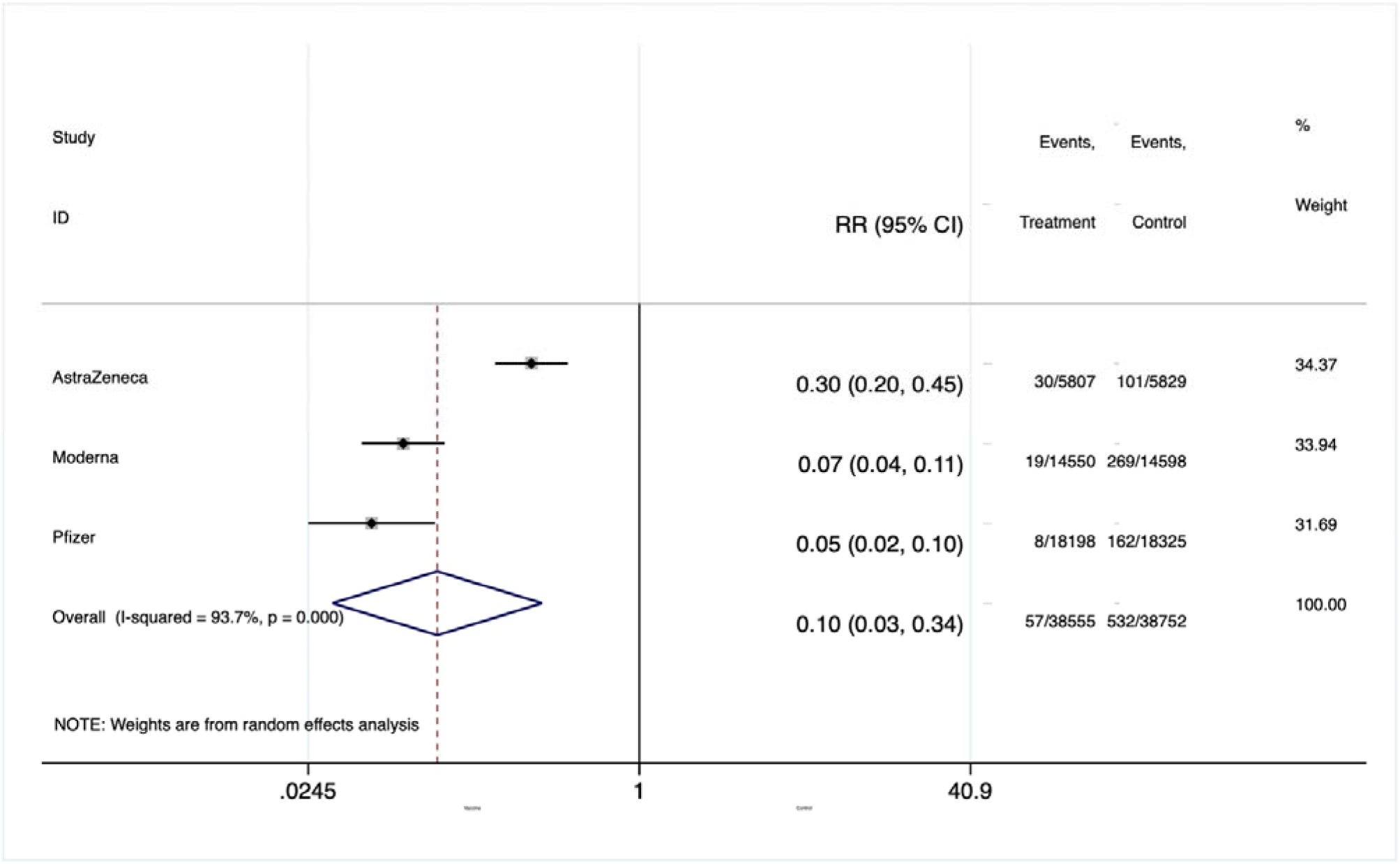
Random effects meta-analysis of COVID-19 vaccines versus control for the prevention of COVID-19 infection.

**Figure 2.**
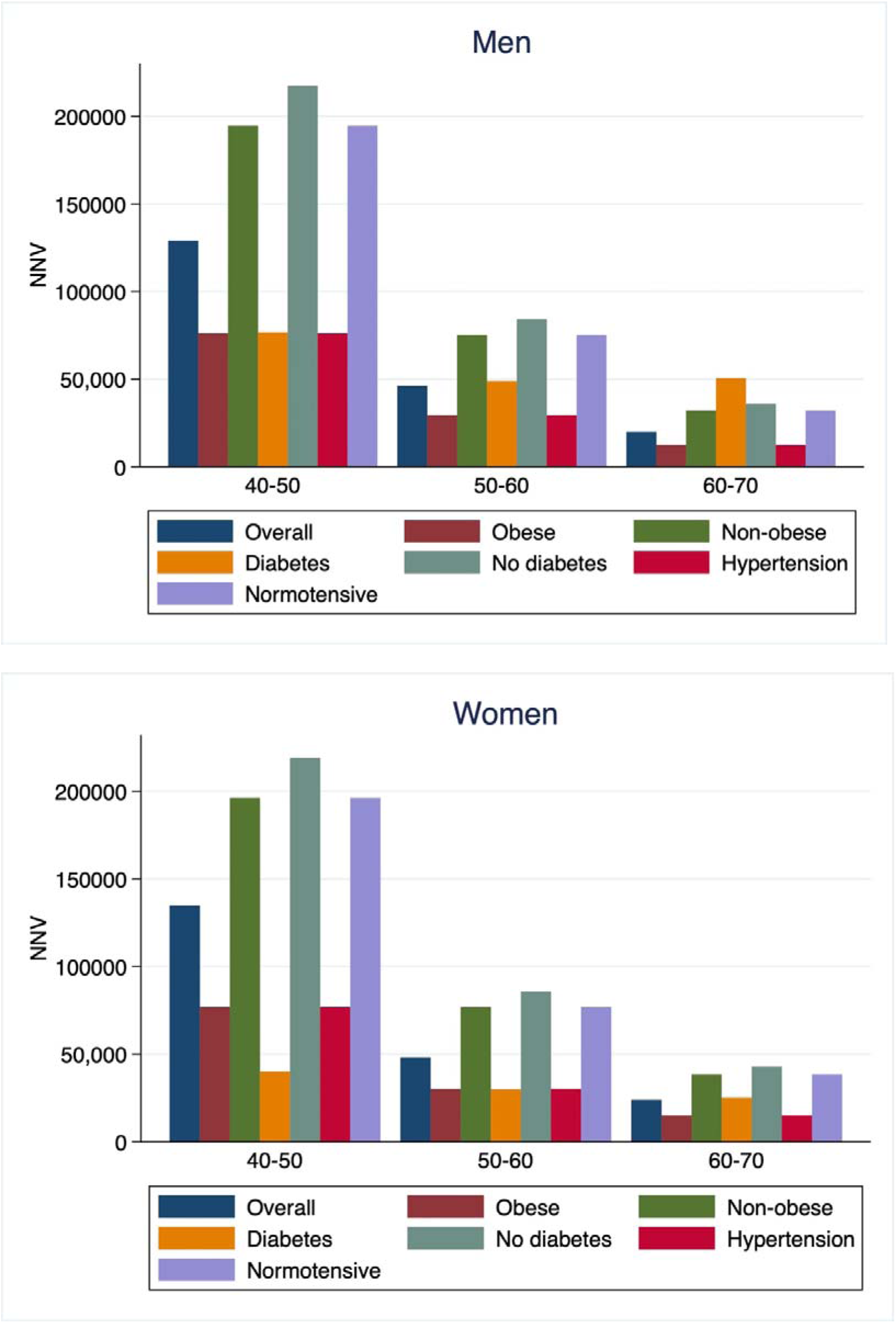
Estimated numbers-needed-to-vaccinate (NNV) to prevent a COVID-19 death. Estimates are stratified by age 40-50 years versus 50-60 years versus 60-70 years.

## DISCUSSION

The major finding from this analysis of Canadian data on middle-aged and older adults is that the number-needed-to-vaccinate to prevent COVID-19 death among those with cardiovascular risk factors (obesity, diabetes or hypertension) is approximately half the number-needed-to-vaccinate among adults without these cardiovascular risk factors. Consequently, the number-needed-to-vaccinate of obese, diabetic and hypertensive adults in any age group is similar to the number-needed-to-vaccinate of non-obese, non-diabetic and normotensive adults 10 years older.

In most regions within Canada, COVID-19 vaccination is offered to the elderly first (along with long-term care residents and frontline healthcare workers), followed by members of the public in cohorts of successively decreasing age. Our analysis supports this approach by demonstrating that vaccinating the elderly first is a highly efficient way of preventing COVID-19 deaths. Once elderly individuals have been vaccinated, there remains uncertainty as to how to prioritize vaccination of the remaining population. It has been recognized that cardiovascular risk factors including obesity, diabetes and hypertension are risk factors for both the acquisition of COVID-19 infection as well as for a fatal outcome in the event of COVID-19 infection. Our analysis, which has been conducted using contemporary data to inform the prevalence of these cardiovascular risk factors in the Canadian population, suggests that preferentially vaccinating individuals with one or more cardiovascular risk factors may be an efficient way to prevent COVID-19 mortality.

This analysis has several limitations. We assumed that COVID-19 vaccines are equally effective among individuals with cardiovascular risk factors as among those without cardiovascular risk factors. While there was no evidence from the randomized, controlled trials of COVID-19 vaccinations to indicate that the efficacy of these vaccines varies according to the recipient’s age, sex or cardiovascular risk factors, data that have yet to be peer-reviewed suggest that among 248 healthcare workers receiving the BNT 162b2 vaccine, the humoral immune response was larger among those with a “normal” body-mass index as compared with a higher body-mass index(12). Also, while we demonstrate a reduction in the number-needed-to-vaccinate to reduce mortality if those with cardiovascular risk factors are vaccinated earlier, we have not estimated the number of quality-adjusted life-years gained by such a strategy. If individuals without cardiovascular risk factors gain more quality-adjusted life-years through COVID-19 vaccination, this may attenuate the apparent advantages of preferentially vaccinating those with cardiovascular risk factors earlier. We have not evaluated the potential impact of preferentially vaccinating adults with chronic diseases other than obesity, diabetes and hypertension because there were limited numbers of these diseases in the PURE data. Public Health England guidance includes diseases such as dementia, kidney disease and recent cancer as conditions warranting earlier vaccination, based in part on data from the UK National Health Service demonstrating individuals with these diseases to be at higher risk of COVID-19 death(13, 14). Lastly, our analysis is based on the Canadian adult population distribution and characteristics. These findings may not be generalizable to populations with substantially different distributions of cardiovascular risk factors, although the general principles are likely to hold.

## BRIEF SUMMARY

- Approximately half as many individuals with a cardiovascular risk factor need to be vaccinated against COVID-19 to prevent related death as compared with individuals without a risk factor.
- Adults with body-mass index ≥30kg/m^2^, diabetes or hypertension should be of a similar priority for COVID-19 vaccination to adults 10 years older with a body-mass index 20 to <30kg/m^2^, no diabetes and no hypertension.

## Data Availability

Summary data are available on request.

https://population.un.org/wpp/Download/Standard/Population/

https://www.canada.ca/content/dam/phac-aspc/documents/services/diseases/2019-novel-coronavirus-infection/surv-covid19-weekly-epi-update-20210212-eng.pdf

https://www.publichealthontario.ca/-/media/documents/ncov/epi/2020/06/covid19-epi-case-identification-age-only-template.pdf?la=en

